# Comparison of a second-generation trabecular bypass (iStent *inject*) to ab interno trabeculectomy (Trabectome) by *exact matching*

**DOI:** 10.1101/2020.01.15.20017582

**Authors:** Yousef Al Yousef, Alicja Strzalkowska, Jost Hillenkamp, André Rosentreter, Nils A. Loewen

## Abstract

**Purpose:** To achieve a highly balanced comparison of trabecular bypass stenting (IS2, iStent inject)with ab interno trabeculectomy (T, Trabectome) by *exact matching*. In a similar study, IS1 (1st generation iStent) had shown a loss of effect at 6 months.

**Methods:** 53 IS2 eyes were matched to 3446 T eyes. Patients were matched using exact matching by baseline IOP, the number of glaucoma medications, and glaucoma type and using nearest neighbor matching by age. Individuals without a close match were excluded. All surgeries were combined with phacoemulsification.

**Results:** A total of 78 eyes (39 in each group) could be matched as exact pairs with a baseline IOP of 18.3±5.1 mmHg and glaucoma medications of 2.7±1.2 in each. IOP in IS2 was reduced to 14.6±4.2 mmHg at 3 months and in T to a minimum of 13.1±3.2 mmHg at 1 month. In IS2, IOP began to rise again at 6 months, eventually exceeding baseline. At 24 months, IOP in IS2 was 18.8±9.0 mmHg and in T 14.2±3.5 mmHg. IS2 had a higher average IOP than T at all postoperative visits (p<0.05 at 1, 12, 18 months). Glaucoma medications decreased to 2.0±1.5 in IS2 and to 1.5±1.4 in T.

**Conclusion:** T resulted in a larger and sustained IOP reduction compared to IS2 where a rebound occurred after six months to slightly above preoperative values. This time course fits bioreactivity data of the IS1.

## Introduction

Following reports of fibrosis [1] and biofilm deposition [2] after trabecular bypass device implantation, we recently compared outcomes of exactly matched pairs of eyes with a first-generation trabecular bypass stent (IS1, iStent, Glaukos Corp., San Clemente, CA) to trabecular ablation (T, Trabectome, MicroSurgical Technology, Redmond, WA) [3]. Such microincisional glaucoma surgeries (MIGS) are now often performed as primary glaucoma surgeries instead of more extensive, traditional procedures (trabeculectomy, tube shunt) because of a low complication rate [4], short procedure time and ability to combine them with outpatient cataract surgery. In eyes with stents, we found evidence of declining function in the form of increasing IOP and glaucoma medications above preoperative levels while IOP and medications reduced in matched eyes that had undergone TM ablation [3]. In the present study, we examined whether a smaller, second-generation trabecular bypass stent (IS2, iStent inject, Glaukos Corp., San Clemente, CA) would be affected differently. Both the IS1 and the IS2 are made out of heparin-coated titanium, but two IS2s are inserted in one session. We compared the IS2 eyes to eyes T using *exact matching*, an advanced statistics method developed for a highly balanced comparison [3, 5, 6]. We hypothesized that this method would again reveal significant effect differences between IS2 and T that can be missed in studies that use group averages [7, 8] and may be caused by chronic responses at that the implant site [9–12].

## Methods

### Study design

The study protocol was approved by the local ethics committee of the University of Würzburg, Germany, and performed in accordance with the ethical standards set forth in the 1964 Declaration of Helsinki and the Health Insurance Portability and Accountability Act. In this study, all patients were included who underwent either IS2 or T between January 2008 and March 2018 in our clinic and associated satellites. An indication for surgery was either a stable IOP with a desire to reduce glaucoma medications at the time of cataract surgery, or an above-target IOP as determined by a glaucoma specialist, while on maximally tolerated topical treatment. To increase the chances of an exact match to the new IS2, of which fewer data exist, data from the Trabectome Study Group database [13, 14] was used to bring the number of T available for an exact match to 3446. Patients younger than 20 years of age, with neovascular or uveitic glaucoma, uncontrolled uveitis, or prior ocular surgery, were excluded. At baseline, patient history was assessed, and data obtained on demographic information, type and stage of glaucoma, best-corrected visual acuity (BCVA), intraocular pressure (IOP), and the number of glaucoma medications. At each follow-up visit, BCVA, IOP, and the number of glaucoma medications were recorded. The primary outcome was a ≥ 20% reduction of IOP compared to baseline or an IOP of less than 21 mmHg to better reflect the most common real-world use pattern of the IS2 as an add-on surgery at the time of cataract surgery rather than a principal glaucoma surgery. Both IS2 and T were combined with phacoemulsification and intraocular lens implantation in all cases. The decision about the continuation of any glaucoma drops was at the discretion of the treating specialist as was the decision to advance to a different glaucoma surgery.

### Statistics

Data were described as frequency, percentage, mean±SD, median, and range. Continuous and categorical variables were compared with the Mann-Whitney U test and chi-squared test. Using exact matching, both groups were matched using preoperative IOP, glaucoma medications, and type of glaucoma, and using *Nearest Neighbor Matchin*g for age [15]. Each unit in group 1 (IS2) was matched using exact matching to all possible control units in group 2 (T). Whereas nearest neighbor matching selected the best matches based on the distance to the value in group 1. P-values of less than 0.05 were considered statistically significant. Mean±SD was used to express continuous variables. Statistical analyses were performed using R [16].

### Surgical technique

IS2 and T were combined with phacoemulsification and lens implantation in all cases. IS2 implantation was done after cataract surgery while T was done first and followed by cataract surgery. IS2 was implanted through a temporal clear corneal incision and in an ophthalmic viscoelastic device stabilized anterior chamber at the conclusion of cataract surgery [17]. Schlemm’s canal was identified by allowing blood from the episcleral veins to reflux in relative hypotony. Under direct gonioscopic view, the tip of the inserter was placed against the nasal TM, and the stents were inserted by piercing through the TM into Schlemm’s canal before releasing. The inserter was retracted, and the viscoelastic removed [17].

T was performed as described before [18]. Briefly, a 1.6 mm uniplanar, temporal clear corneal incision was created. Under direct gonioscopic visualization, the tip of the handpiece was inserted into Schlemm’s canal, and the TM was ablated counterclockwise, followed by clockwise ablation with a total length of around 120° [19–21]. Ablation was started with the power set to 0.8 mW and increased as necessary. The handpiece was withdrawn from the anterior chamber. T was done before cataract surgery to provide the highest corneal transparency for angle surgery and because this incision is smaller than the one needed for cataract surgery. After T, a viscoelastic device was injected to form the anterior chamber immediately before enlarging the clear corneal incision for cataract surgery.

In both IS2 and T, postoperative treatment consisted of a combination of a topical antibiotic for one week and a steroid tapered over four weeks. Glaucoma medications were stopped on the day of surgery and restarted as needed.

## Results

Of the 53 IS2 eyes, 39 could be matched to T to create 39 near-identical pairs. Due to *exact matching*, there was no difference in IOP, the number of IOP-lowering medications, glaucoma type, or VF loss between groups (p>0.05). **Table 1** shows the baseline characteristics of each group, both of which were combined with cataract surgery. IS2 and T had the same preoperative IOP of 18.3±5.1 mmHg. In IS2, an IOP trough of 14.6±4.2 mmHg was reached at three months (p=0.04 compared to preoperative). Remarkably, IOP then continued to slowly increase from six months onward throughout the remainder of the study, eventually reaching the baseline average (p>0.05, **Figure 1A)**. In T, IOP was reduced to a minimum of 13.1±3.2 mmHg at one month (p=0.01, **Figure 1A)**. It stayed at that level throughout the observation time (p<0.02 all time points compared to preoperative). At 24 months, IOP in IS2 was 18.8±9.0 mmHg while in T, it was 14.2±3.5 mmHg. T had a lower average IOP than IS2 at all postoperative visits (p<0.05 at 1, 12, 18 months, **Table 2**).

**Table 1.**
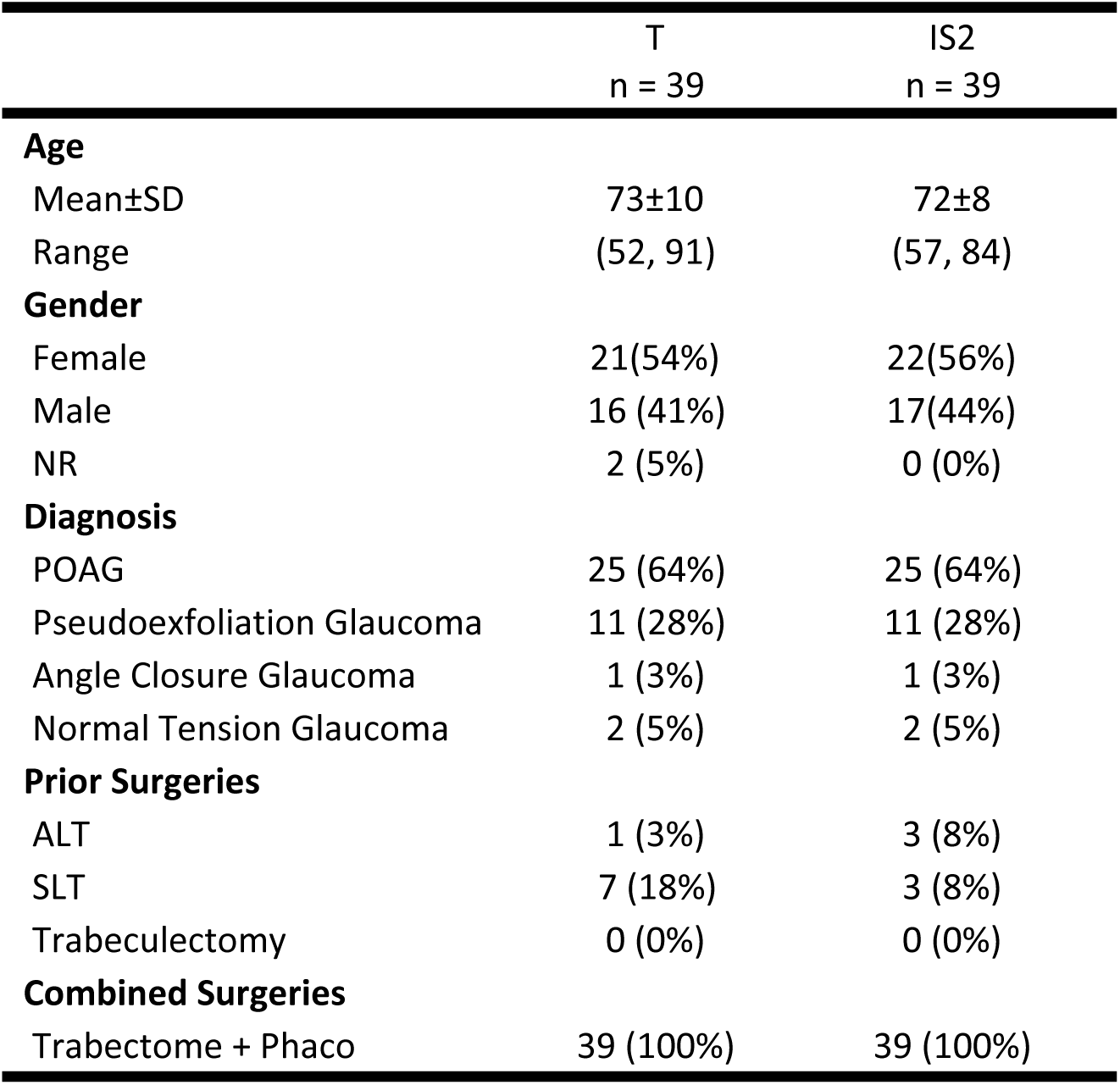
Demographics.

|  | T<br>n = 39 | IS2<br>n = 39 |
| --- | --- | --- |
| **Age** |  |  |
| Mean±SD | 73±10 | 72±8 |
| Range | (52, 91) | (57, 84) |
| **Gender** |  |  |
| Female | 21(54%) | 22(56%) |
| Male | 16 (41%) | 17(44%) |
| NR | 2 (5%) | 0 (0%) |
| **Diagnosis** |  |  |
| POAG | 25 (64%) | 25 (64%) |
| Pseudoexfoliation Glaucoma | 11 (28%) | 11 (28%) |
| Angle Closure Glaucoma | 1 (3%) | 1 (3%) |
| Normal Tension Glaucoma | 2 (5%) | 2 (5%) |
| **Prior Surgeries** |  |  |
| ALT | 1 (3%) | 3 (8%) |
| SLT | 7 (18%) | 3 (8%) |
| Trabeculectomy | 0 (0%) | 0 (0%) |
| **Combined Surgeries** |  |  |
| Trabectome + Phaco | 39 (100%) | 39 (100%) |

**Table 2.** Mean IOP and number of Medication for Trabectome and IStent group at each follow-up time point (Welch two-sample t-test, significance level set at ≤0.05).

| Time | IOP |  |  | Rx |  |  |
| --- | --- | --- | --- | --- | --- | --- |
|  | Mean±SD |  | P-value | Mean±SD |  | P-value |
|  | T | IS2 |  | T | IS2 |  |
| Baseline | 18.3±5.1 | 18.3±5.1 |  | 2.7±1.2 | 2.7±1.2 |  |
| 1 month | 13.1±3.2 | 15.5±4.7 | <0.01* | 2.3±1.6 | 1.9±1.5 | 0.89 |
| 3 months | 13.2±3.3 | 14.6±4.2 | 0.05 | 2.1±1.6 | 1.7±1.4 | 0.87 |
| 6 months | 14.0±3.2 | 14.7±2.8 | 0.17 | 1.8±1.3 | 1.8±1.5 | 0.51 |
| 12 months | 13.3±2.5 | 16.5±5.2 | 0.01* | 1.6±1.3 | 1.9±1.5 | 0.16 |
| 18 months | 14.2±2.7 | 17.3±6.8 | 0.04* | 1.8±1.4 | 2.0±1.5 | 0.27 |
| 24 months | 14.2±3.5 | 18.8±9.0 | 0.10 | 1.5±1.4 | 2.0±1.5 | 0.11 |

**Figure 1:**
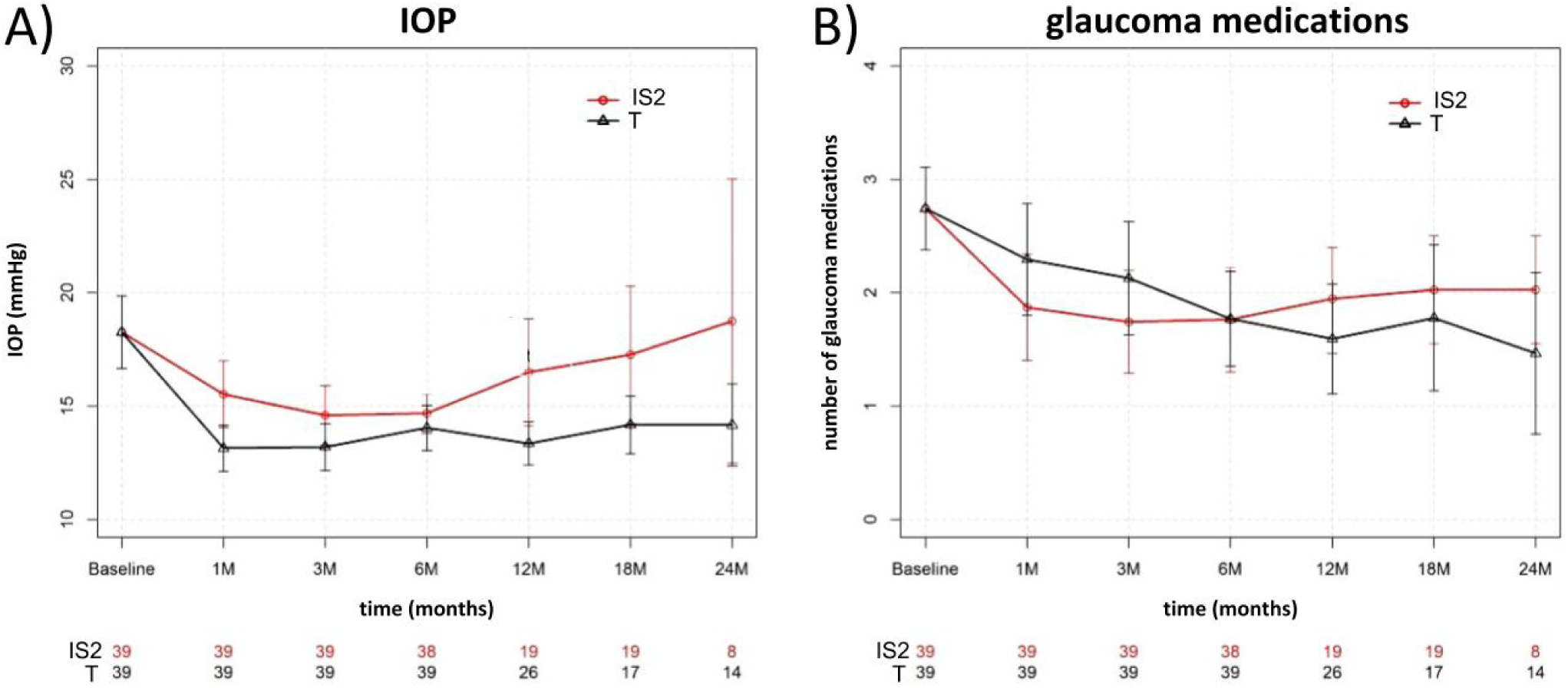
Mean IOP and number of glaucoma medications for IS2 (iStent *inject*) and T (trabectome). A) IOP decreased in IS2 through month 3 before starting to rise. In T, IOP decreased through month 1 and remained at this level throughout the remainder of the study. B) Glaucoma medications decreased in IS2 before starting to rise again after month 6. In T, medications decreased throughout the study (mean±SD; subject count at each time point).

Glaucoma medications started at a matched value of 2.7±1.2 in IS2 and T. In IS2, they decreased to 1.7±1.4 by month three (p=0.04), only to increase to a slightly higher average starting at six months, becoming not significantly different from preoperative counts by month 12 (p>0.05) but apparently not enough to prevent the IOP from rising. At 24 months, the medication count in IS2 was 2.0±1.5 (p>0.05 compared to preoperative, **Figure 1B)**. In T, medications decreased from the exactly matched count of 2.7±1.2 at baseline, becoming significantly lower at six months (1.6±1.3, p=0.03) and declining to 1.5±1.4 at 24 months (p=0.04, **Figure 1B)**.

Using a definition of success commonly applied to trabecular bypass stents, a final IOP of ≤ 21 mmHg or a 20% IOP reduction from baseline, 97% of T and 95% of IS2 achieved this goal (**Figure 2**). A second surgery was required in one patient in T and in two in IS2. No intra- or postoperative vision-threatening complications such as choroidal effusion, sustained hypotony, choroidal hemorrhage, or infection occurred.

**Figure 2:**
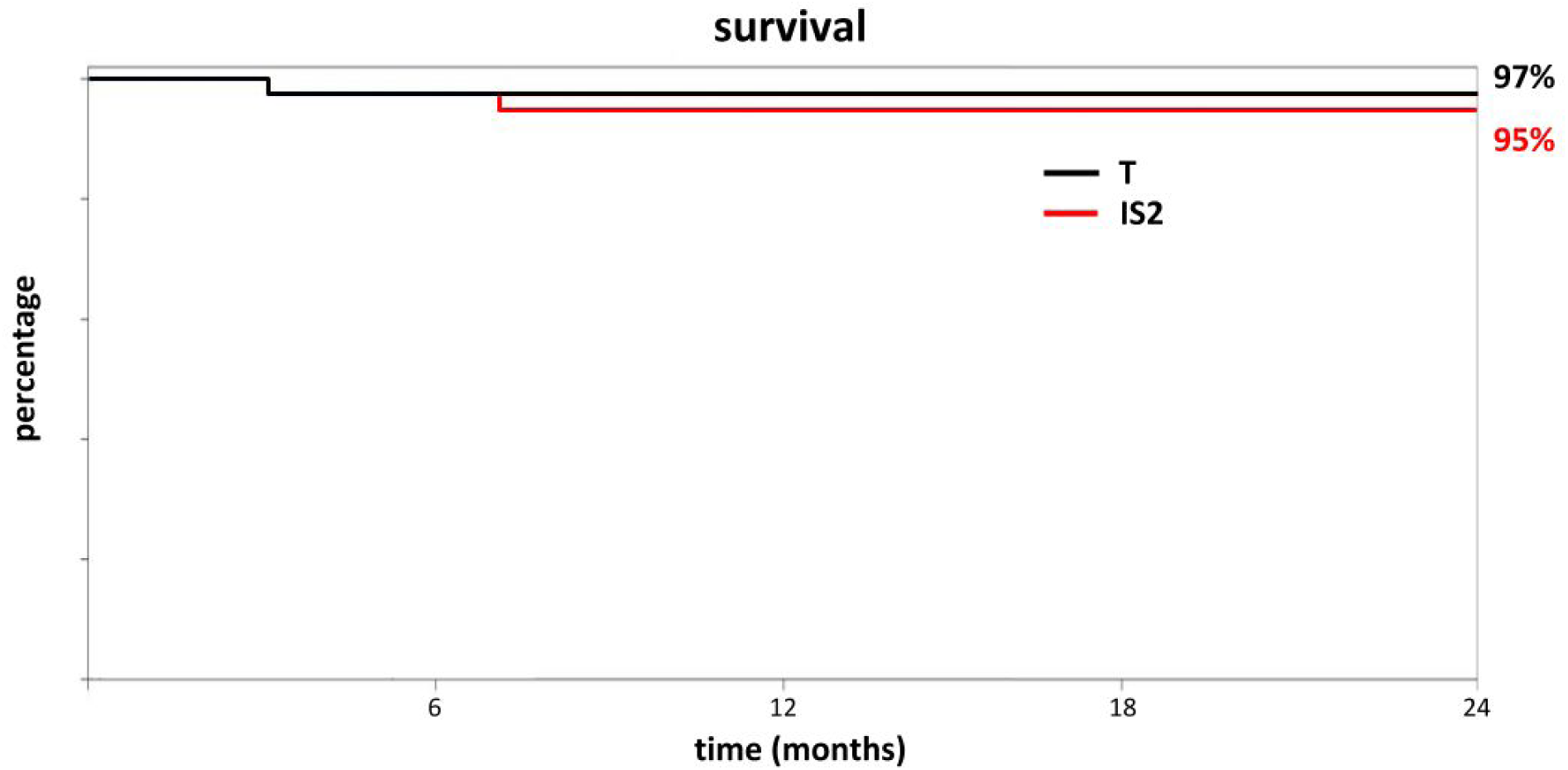
Procedure survival curve for IS2 and T with 24 months of follow-up time indicates a relatively high success rate despite a modest IOP and medication reduction. The criteria were a ≥ 20% IOP reduction *or* an IOP of less than 21 mmHg reflecting a common use pattern of IS2 as an add-on surgery at the time of cataract surgery rather than a principal glaucoma surgery.

## Discussion

The study presented here applied exact matching to detect essential differences between IS2 and T using real-world data. Such differences can be missed in studies that use nonrandomized patient assignments or apply simpler statistics [8, 22]. IS2 is a second-generation trabecular bypass stent [17] made out of the same material as the IS1 (iStent, Glaukos Corp., San Clemente, CA) but considerably smaller [23], allowing to fit two into one injector. The IS2 is implanted using a TM-puncturing forward movement, simplifying the technique and doubling the chance to correctly place the device in Schlemm’s canal. In contrast, T eyes do not retain an implant and the TM is molecularized using plasma [18, 24].

In the IS2 patients followed here, we observed a slow loss of IOP reduction after six months that was absent in T. The average IOP eventually exceeded the preoperative IOP by 24 months and was indistinguishable from the preoperative IOP, likely not reaching significance due to a decreased subject count. In contrast, T continued at an approximately 20% reduced IOP through the end of the study.

Medications in IS2 were increased slightly around six months but did not experience a rebound above the preoperative level. In T, medications continued to decline throughout the study. Our experience with the IS2 resembled the IS1 [3] except that IS1 patients had an IOP increase above baseline that was higher (about 15%) despite counteracting this trend by using approximately 30% more glaucoma medications compared to the companion T eyes.

IS2 and IS1 are safe add-on procedures that were approved in conjunction with cataract surgery [17, 25], and this use pattern continues to be the most common one. In contrast, T is used across a spectrum of glaucoma severity [26, 27], after failed glaucoma surgery [14, 28] on its own [26] or combined with cataract surgery [27]. The survival rates we computed imply a high clinical success rate for both IS2 and T, but they are based on use criteria common for MIGS with an IOP below 21 mmHg or an IOP reduction by 20%. The clinical utility of a medication reduction by 0.7 as obtained here with the IS2 depends on the motivation and goal of the affected individual. The IS2 IOP and medication change observed here might be caused by fibrosis [1] and biofilm deposition described previously [2]. Although one might expect a more substantial effect from implanting two trabecular bypass IS2 stents than one IS1, the smaller aperture of the IS2 may make it more vulnerable to TM reactivity [9–12].

The lowest IOP any TM bypass or ablation surgery could theoretically achieve is limited to a pressure of about 8 mmHg that is present in the episcleral venous veins [29]. This limit avoids an excessive pressure reduction that can result in hypotony in traditional glaucoma surgery [30] and after MIGS that drain into the suprachoroidal [31] or subconjunctival space [32]. It remains unclear why trabecular bypass, disruption, or ablation methods do not routinely achieve a postoperative IOP around 8 mmHg, but post-trabecular resistor elements that have been observed experimentally might play a role [33, 34].

Limitations of our study are that it was designed to detect differences between IS2 and T that cannot readily be discovered otherwise. As a result, it can only inform about intragroup changes within limits. For instance, patients with a high preoperative IOP undergoing T typically have a much more substantial IOP reduction than those with a low preoperative IOP, as examined here [26, 27]. All patients also had phacoemulsification, which can lower IOP on its own [25]. It has been speculated that the IOP reduction after phacoemulsification is due to a trabeculoplasty-like reaction [3, 35]. IS2 might have been inadvertently helped by this additional effect [36]. However, there is no significant contribution from cataract surgery to IOP reduction in T as our prior studies found [37, 38]. Presumably, this is the case because most of the conventional outflow after T occurs through the 120 to 180 degrees of nasal, unroofed Schlemm’s canal [39]. In IS2, the nasal Schlemm’s canal remains covered by glaucomatous TM and can be affected by a trabeculoplasty-like effect of cataract surgery.

In summary, a highly balanced comparison of IS2 and T by exact matching showed that reduction of IOP and the number of glaucoma medications is lower and sustained in T. IOP in IS2 rebound 6 to 24 months after surgery, eventually reaching preoperative levels.

## Data Availability

All data presented is contained in the manuscript

